# RNA-extraction-free diagnostic method to detect SARS-CoV-2: an assessment from two States, India

**DOI:** 10.1101/2021.09.19.21263807

**Authors:** Madhumathi Jayaprakasam, Sumit Aggarwal, Arati Mane, Vandana Saxena, Amrita Rao, Bhaswati Bandopadhyay, Banya Chakraborty, Subhasish Kamal Guha, Mekhala Taraphdar, Alisha Acharya, Bishal Gupta, Sonia Deb, Aparna Chowdhury, Kh Jitenkumar Singh, Prashant Tapase, Ravindra M Pandey, Balram Bhargava, Samiran Panda

**Affiliations:** Division of Epidemiology and Communicable Diseases, Indian Council of Medical research (ICMR), Ansari Nagar, New Delhi, India; Department of Microbiology, ICMR- National AIDS Research Institute (NARI), Pune, India; Department of Microbiology, School of Tropical Medicine, Kolkata, India; ICMR-National Institute of Medical Statistics (NIMS), Ansari Nagar, India; Department of Biostatistics, All India Institute of Medical Sciences (AIIMS), New Delhi, India

**Keywords:** SARS CoV-2, RNA-extraction free method, Diagnostic test, Heat-inactivation, Dry swab, Performance-evaluation

## Abstract

With increasing demand for large numbers of testing during COVID-19 pandemic, came alternative protocols with shortened turn-around time. We evaluated the performance of such an approach wherein 1138 consecutive clinic attendees were enrolled; 584 and 554 respectively from two independent study sites in the cities of Pune and Kolkata. Paired nasopharyngeal and oropharyngeal swabs were tested by using both reference and index methods in blinded fashion. Prior to conducting RT-PCR, swabs collected in viral transport medium (VTM) were processed for RNA extraction (reference method) and swabs collected in dry tube without VTM were incubated in Tris-EDTA-Proteinase K buffer for 30 minutes and heat inactivated at 98°C for 6 minutes (index method). Overall sensitivity and specificity of the index method were 78.9% (95% CI 71% to 86%) and 99 % (95% CI 98% to 99.6%) respectively. Agreement between the index and reference method was 96.8 % (k = 0.83, SE=0.030). The reference method exhibited enhanced detection of viral genes (E, N and RdRP) with lower Ct values compared to the index method. The index method can be used for detecting SARS-CoV-2 infection with appropriately chosen primer-probe set and heat treatment approach in pressing time; low sensitivity constrains its potential wider use.

## Introduction

The COVID-19 pandemic swept through the world with unprecedented speed and impact on lives and livelihoods [1]. Within four months of its onset, more than 118,000 cases and 4,291 deaths were reported from 114 countries. All of these happened following an outbreak of ‘unusual cases of pneumonia’ notified for the first time from the Wuhan city of Hubei province, China in December 2019 [2]. Such a rapid spread of the causative virus SARS-CoV-2 reminded humankind of the influenza pandemic causing havoc about 100 year ago [3,4]. Developing simple and reliable diagnostic tests appeared paramount in this context as care service related needs escalated and demand for tools to conduct quick screening and survey also increased [5].

As with many other infectious diseases, SARS-CoV-2 infection is detected reliably by the real-time polymerase chain reaction (RT-PCR) as it is a highly sensitive and specific tool [6]. While the Center for Disease Control (CDC), USA recommended the gene targets for two nucleocapsid proteins (N1 and N2) of SARS-CoV-2 for diagnostic assays [7], the World Health Organization (WHO) proposed using envelope (E) gene target for first line screening and RNA dependent RNA Polymerase (RdRP) for confirmation [8,9]. Notably, assays using E and N2 gene primers were found to be more sensitive [10]. The combination of two gene targets is recommended to enhance accuracy of diagnosis in the context of possible viral genetic variability; one from the conserved region of the virus and another from SARS CoV-2 specific region of the genome [8].

Several alternative protocols described ways to simplify RT-PCR test by excluding the RNA extraction step [11-14]. These modifications attempted to reduce the turn-around time for quickly obtaining test results and also to address the issue of shortage of RNA extraction kits when the demand runs high. Heating of naso-pharyngeal swab specimens in transport medium and skipping RNA extraction step before proceeding to conduct RT-PCR has been reported to be fast and reliable [12]. Direct heating of viral extracts from swab specimens for 5 minutes at 98 °C resulted in 97% sensitivity and 100% specificity when examined against purified RNA as gold standard [15]. Direct RT-PCR assay with heat-inactivated or lysed samples using generic buffers like Tris or Tris-EDTA (TE) served as an effective alternative method [16]. A similar approach to RT-PCR, using heat inactivated TE buffer extract of nasopharyngeal swabs transported in dry tube from sample collection site to the laboratory, has been described from India as well [17]. However, utility of this method and modified version of it as suggested by the Centre for Cellular and Molecular Biology, Hyderabad, India in real world program setting was not examined. This modification was in line with the work of Chu et al [13] for SARS CoV-2 and de Paula et al., [18] for Hepatitis A virus where proteinase K was used along with TE buffer. We assessed the performance of modified version of the test approach of Kiran et al., in program setting for diagnosis of COVID-19, using E, RdRP and N primer-probe based assay.

## Methods

The current investigation took place during 10^th^ November through 11^th^ December 2020. The proposal for evaluation was developed in early October 2020 and approval was obtained from the Central Ethics Committee for Human Research (CECHR) of Indian Council of Medical Research (ICMR) on 30^th^ October 2020. Written informed consent was obtained from individuals consenting to participate in this study.

### Study settings and participants

The present investigation was conducted at two sites in India namely, the ICMR-National AIDS Research Institute (ICMR-NARI), Pune in the western state of Maharashtra and the School of Tropical Medicine (STM), Kolkata in the eastern state of West Bengal. Necessary approvals were obtained from the Ethics Committees of these two respective institutes as well. Consecutive clinic attendees (≥ 18 year of age) at the designated study sites, who came for SARS-CoV-2 testing, were invited to participate in this investigation.

### Implementation

Each consenting clinic attendee was registered on a web-based portal maintained by ICMR with a specimen referral form (SRF) number created at the collection site, which was used for labeling the viral transport medium (VTM) containing tube. In order to ensure blinding, a different set of unique codes were randomly generated from the ICMR-headquarter, New Delhi for each study site using Excel based tool for labelling the corresponding swabs collected and placed in dry tubes. The link page, containing ICMR SRF number and the paired unique code for swabs in dry tubes for each enrolled participant, was available only with the designated staff at the respective study sites. Blinding was ensured through barring of the laboratory staff involved in test procedures and generation of test results to the link page.

### Sample collection and processing

‘Specimen Collection, Packaging and Transport Guidelines for 2019 Novel Coronavirus (2019-nCoV)’, was adhered to during study implementation [19]. Two naso-pharyngeal swabs and two oro-pharyngeal swabs were collected from each of the enrolled participants in single sitting. The swabs (one naso-pharyngeal and one oro-pharyngeal) saved in labelled VTM tubes and those kept in labelled dry tubes, were transported to the participating laboratories and processed on the same day of sample collection.

### Reference test and Index test

i. *RT-PCR test with swab specimens transported in VTM tube (reference method):* The reference method used one nasopharyngeal and one oropharyngeal swab (HiMedia™ Laboratories Pvt Ltd, Mumbai, India) collected from each participant and saved in the tube containing 3 ml VTM (HiMedia™ Laboratories Pvt Ltd, Mumbai, India). About 200 µl of the VTM extract was used for RNA extraction followed by RT-PCR assay [9].
ii. *Dry swab-based RT-PCR (index method):* One nasopharyngeal swab and one oropharyngeal swab collected from each participant were transported to the laboratories in 10 ml sample collection tubes (HiMedia™ Laboratories Pvt Ltd, Mumbai, India) without adding VTM to them. At the laboratories 400 µl of Tris-EDTA-Proteinase K (TE-P) buffer [10mM Tris (pH 7.4), 0.1mM EDTA, and 2 mg/ml Proteinase K] (Bio Ultra, for molecular biology, Sigma-Aldrich, Bangalore, India) was added to swab specimens transported in dry tube and incubated for 30 minutes at room temperature. About 50 µl of the TE-P extract was aliquoted into PCR tube and was heat inactivated at 98°C for 6 minutes using thermal cycler (ICMR-NARI site) or heat block (STM site). The heat inactivated extract was then used as template for RT-PCR reaction (Figure 1).

**Figure 1.**
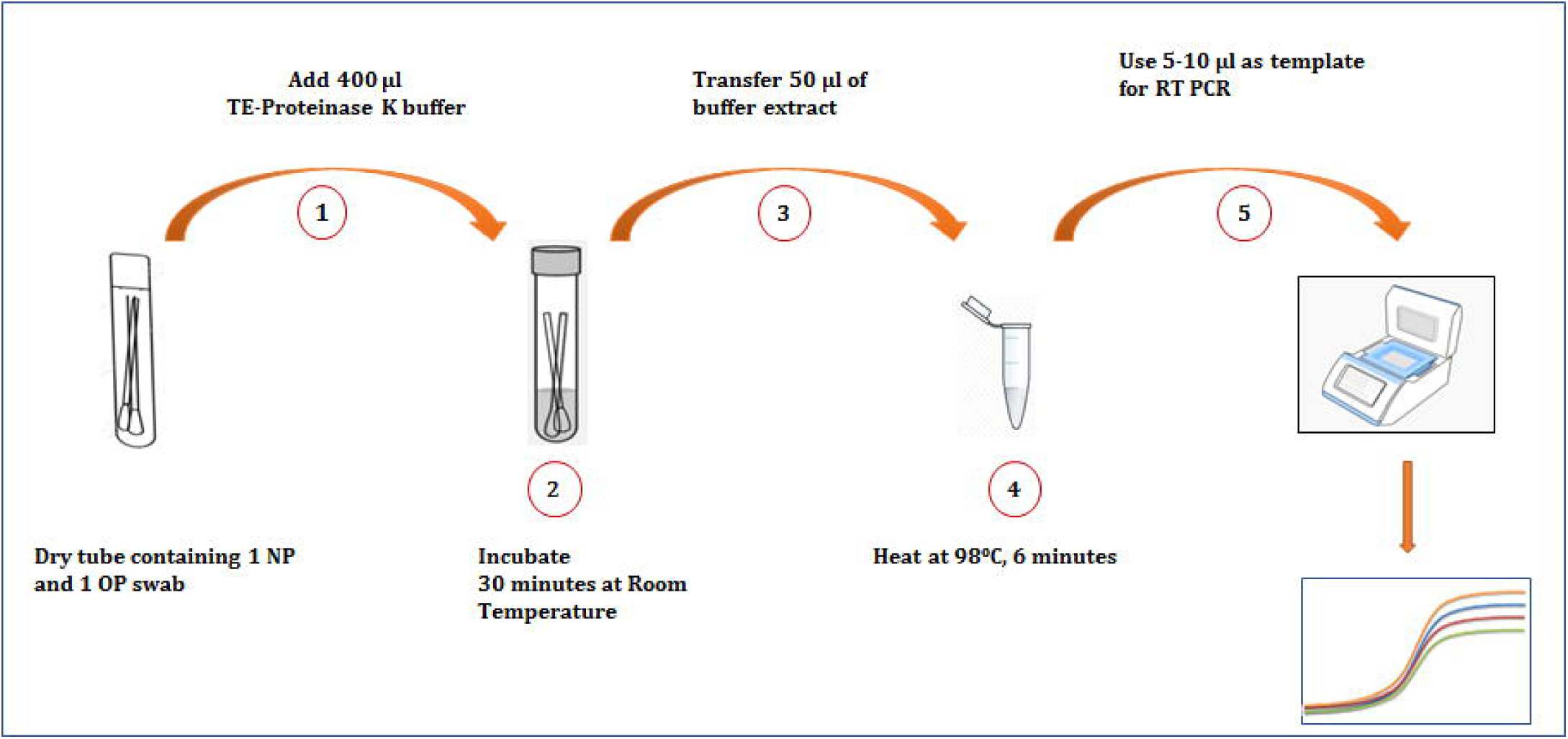
Schematic illustration of modified dry tube-based heat inactivation method followed by RT-PCR for detection of SARS-CoV-2. 1. Addition of TE-Proteinase K buffer to the tube containing swabs 2.Incubation of swabs in buffer to extract viral particles 3. Transfer of viral extract 4. Inactivation of the virus by heating. 5. Setting up RT-PCR reaction and interpretation of assay. NP= Nasopharyngeal swab, OP= Oropharyngeal swab.

### Nucleic Acid Amplification Assay

RNA extraction from clinical specimens transported in VTM tubes was carried out as per instructions accompanying the commercial RNA extraction kit (QIAmp viral RNA Mini Kit, QIAGEN, New Delhi, India). The RT-PCR reaction was carried out using ‘CoviDx mPlex-4R SARS-CoV-2’ (NeoDx Biotech Labs Private Limited, DSS Imagetech, New Delhi, India) as per manufacturer’s instructions with primer-probe sets (Table 1) for detection of the SARS-CoV-2 specific genes E, N and RdRP. Human RNase P was used as internal control in this single tube assay. Briefly, a 25 µl reaction was set up containing 8 µl template (purified RNA from VTM sample for reference method and heat inactivated dry swab lysate for index method), 12.5 µl of 2X master mix, 1.25 µl 20X primer and probe mix and 3.25 µl nuclease free water. In each assay a positive control and no template control (NTC) were included. The RT-PCR assays were conducted on ‘CFX96-IVD Real-time PCR system’ (Bio-Rad Laboratories India Pvt. Ltd., Gurugram, Haryana, India) using the following cycling conditions; 50 °C for 15 minutes for reverse transcription, 95 °C for 2 minutes, and 45 cycles of 95 °C for 15 s and 58 °C for 30 s.

**Table 1.**
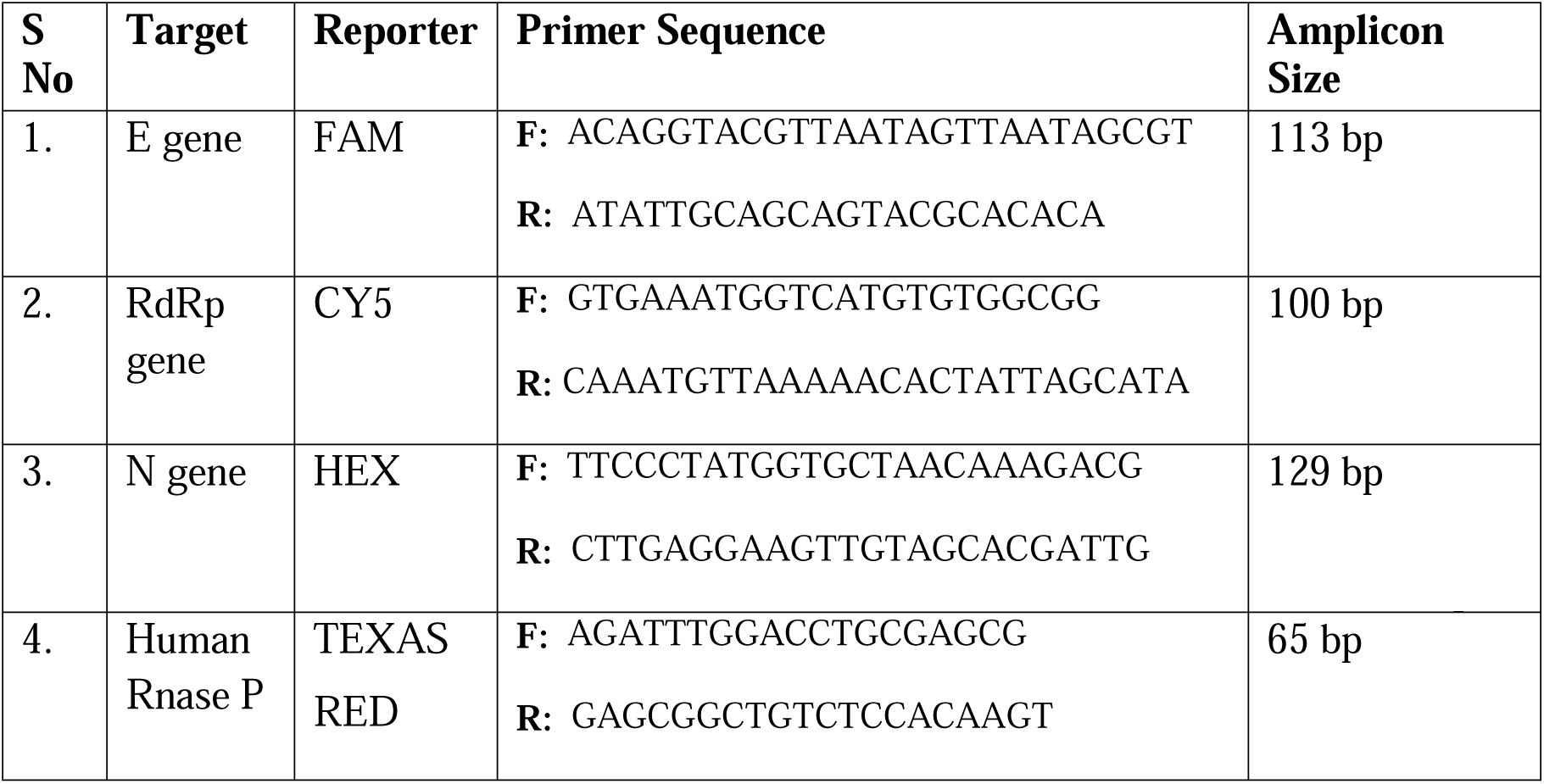
Primer Probe sets of RT-PCR Kit used for assay

### Sample size estimation and data analysis

An earlier evaluation of the RNA-extraction-free dry swab-based RT-PCR method [17] in clinic setting was conducted by us and estimated to have 56% sensitivity and 95% specificity [20]. The modified index method (described above) was expected to have improved sensitivity and thus we conservatively assumed it to be 75% with minimum acceptance lower confidence limit of 60% based on which the calculated number of cases required was 107 [21]. With the recorded prevalence of 20 % SARS-CoV-2 infection among clinic attendees in Pune and Kolkata during the current study, the number of SARS-CoV-2 negative individuals to be included was calculated as 428 {107 x (1-0.2/0.2)} = 428; the total estimated sample size being 535.

A cycle threshold (Ct) value of 40 or less was considered as positive. The binary outcome (yes/no), in the form of presence or absence of SARS-CoV-2 infection generated by the index method was assessed against the results obtained following RT-PCR tests on swab specimens transported in VTM. The sensitivity, specificity, concordance, discordance, positive predictive value, negative predictive value and agreement between the tests and their 95% confidence interval (CI) were computed using Stata version 13.1 (StataCorp LP, College Station, TX, USA). Graphpad Prism (version 5) and R-CRAN (version 4.0.3) with ggplot2 library was used for graphical representations.

## Results

### Participants

Consecutive clinic attendees at the two study sites were enrolled. While 15 of the 600 (2.5%) attendees at ICMR-NARI, Pune site refused to provide consent, 96 of the 650 attendees at STM, Kolkata (14.8%) did so. Information obtained from 584 participants by ICMR-NARI (one specimen could not be analyzed due to inadequate volume) and 554 participants by STM, Kolkata were included in the analyses. Each site thus fulfilled sample size requirement on its own and allowed examination of performance of the index test in two different real world settings independent of each other and thus fulfilling the criteria of conducting evaluation in different settings.

The majority of the participants were male (767/1138; 67%); age ranging from 18 to 85 year (Table 2). Nearly 30% of the participants were symptomatic (342/1138); most common ones being fever (52%), cough (35%), bodyache (12%), sore throat (7%), breathlessness (3.5%) and anosmia (3.5%).

**Table 2.** Demographic profile of study participants

| Characteristics | Frequency (%) |
| --- | --- |
| Total number of participants | 1138 |
| **Male (n=767)** |  |
| Mean age ± SD (year) | 38.47 ± 13.2 |
| Median age (year) [IQR] | 37 [27-49] |
| **Female (n=371)** |  |
| Mean age ± SD (year) | 35.97 ± 13.7 |
| Median age (year) [IQR] | 34 [24-45] |
| **Age group (year)** | **Total n (%)** |
| 18-30 | 437 (38) |
| 31- 40 | 247 (22) |
| 41-50 | 235 (21) |
| 51-60 | 160 (14) |
| >60 | 59 (5) |
| Total | 1138 (100) |
| **Symptoms (n=342)** |  |
| Cough | 120 (35.1) |
| Fever | 178 (52.0) |
| Bodyache | 40 (11.7) |
| Breathlessness | 12 (3.5) |
| Sore throat | 24 (7.0) |
| Nausea | 4 (1.2) |
| Heamoptysis | 1 (0.3) |
| Vomiting | 1 (0.3) |
| Chest pain | 2 (0.6) |
| Anosmia | 12 (3.5) |
| Others | 79 (23.1) |

### Comparison of Ct values - reference vs index method

Heat-maps of Ct values for E, RdRP and N genes detected by reference and index methods were plotted. Samples, which were detected having at least one of these genes by VTM-based method, were used for comparisons and were examined to explore how did the index method perform against them. The N gene primer-probe set showed superior performance compared to the other two genes by both reference and index methods. Figure 2 presents comparative data visualization with juxtaposition pertaining to the three aforementioned genes along with the internal control (human Rnase P). The reference method could detect either one of the three genes (E, RdRP or N gene) in 54 samples at ICMR-NARI and 71 samples at STM. However, the index method could detect either one of the three genes in 45 out of the aforementioned 54 samples at ICMR-NARI and 55 of the 71 samples at STM (Figure 2a & 2b). The index method could not detect any of the three target genes in 17 % (9/54) of the clinical specimens at ICMR-NARI and the proportion of such missed events had risen to 23 % (16/71) at STM. Parity between the reference and extraction free methods in terms of detecting positive specimens was better at ICMR-NARI (45/54; 83.3 %) (Fig 2a) compared to the results obtained at STM (55/71; 77.5 %) (Fig 2b). This difference could be explained by the difference in heat treatment methods used by the respective centres. While the STM site used heat block for maintaining 98°C at 6 minutes, the ICMR-NARI site had used thermal cycler.

**Figure 2.**
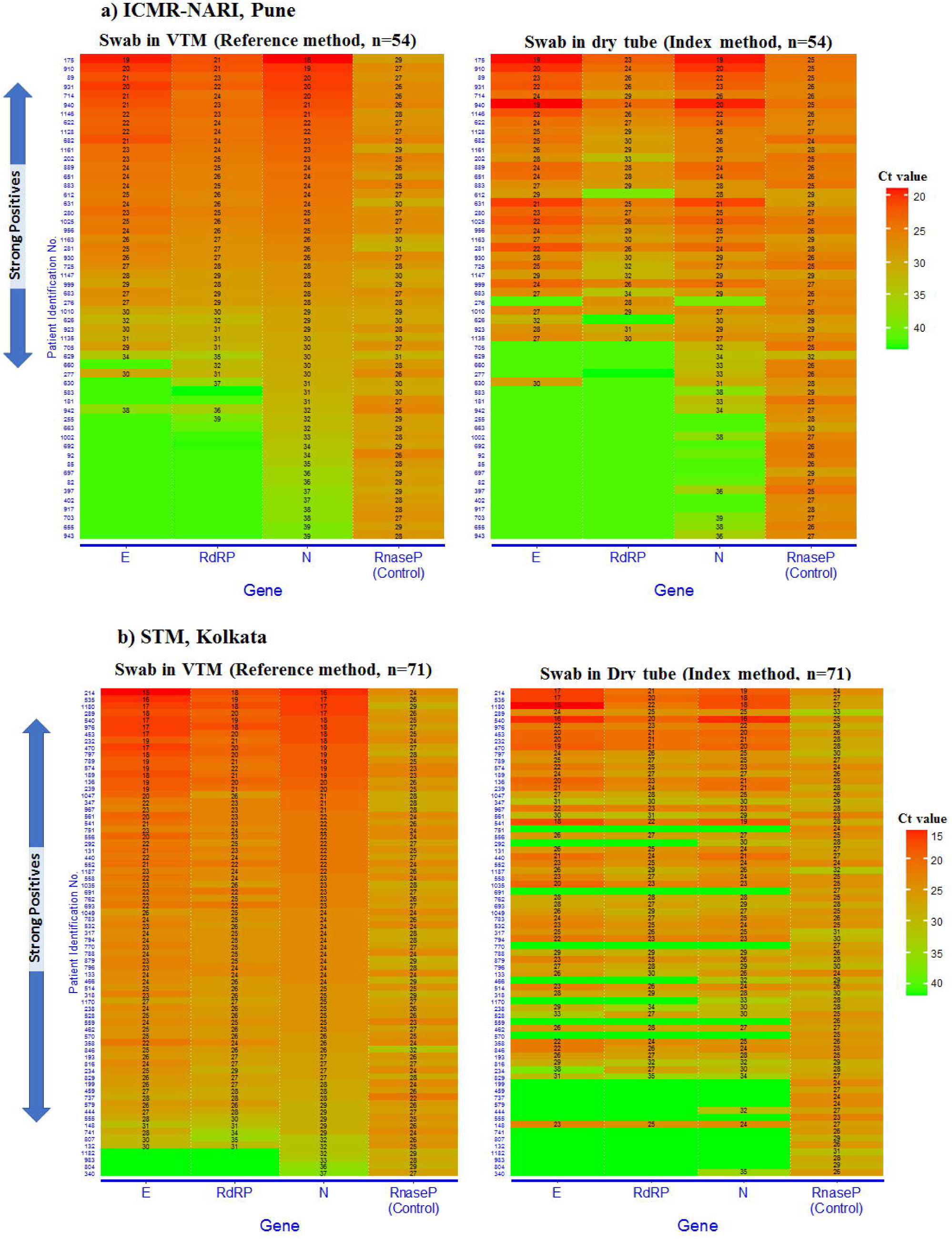
Heat map of cycle threshold (Ct) values for E, RdRP and N genes detected by reference and index methods on clinical samples from ICMR-NARI (n=54) and STM, Kolkata (n=71). The heat map is ranked by N gene Ct. Ct values of human Rnase P used as control in RT PCR is shown in the right. A Ct value ≤ 40 is considered as positive. Samples that are positive for all three viral genes are indicated as strong positives by an arrow on the left.

### Distribution of Ct values

We compared matched Ct values generated by both reference as well as index method. Samples which were tested positive by reference method for each gene were used for the analysis of Ct values. Reference method-based RT-PCR results had 1-10 Ct values lesser than those generated by index method for E, RdRP and N genes in more than two-third of the samples (Figure 3). The mean Ct values (± SD) for target genes detected by reference method were as follows; E= 23.69 ± 4.03, RdRP = 25.59 ± 4.06 and N = 25.76 ± 5.33. These values were significantly lower (p<0.0001, Wilcoxon signed rank test), compared to the values generated by the dry swab method (E = 24.42 ± 4.01, RdRP = 26.80 ± 3.32 and N = 26.61 ± 4.87).

**Figure 3.**
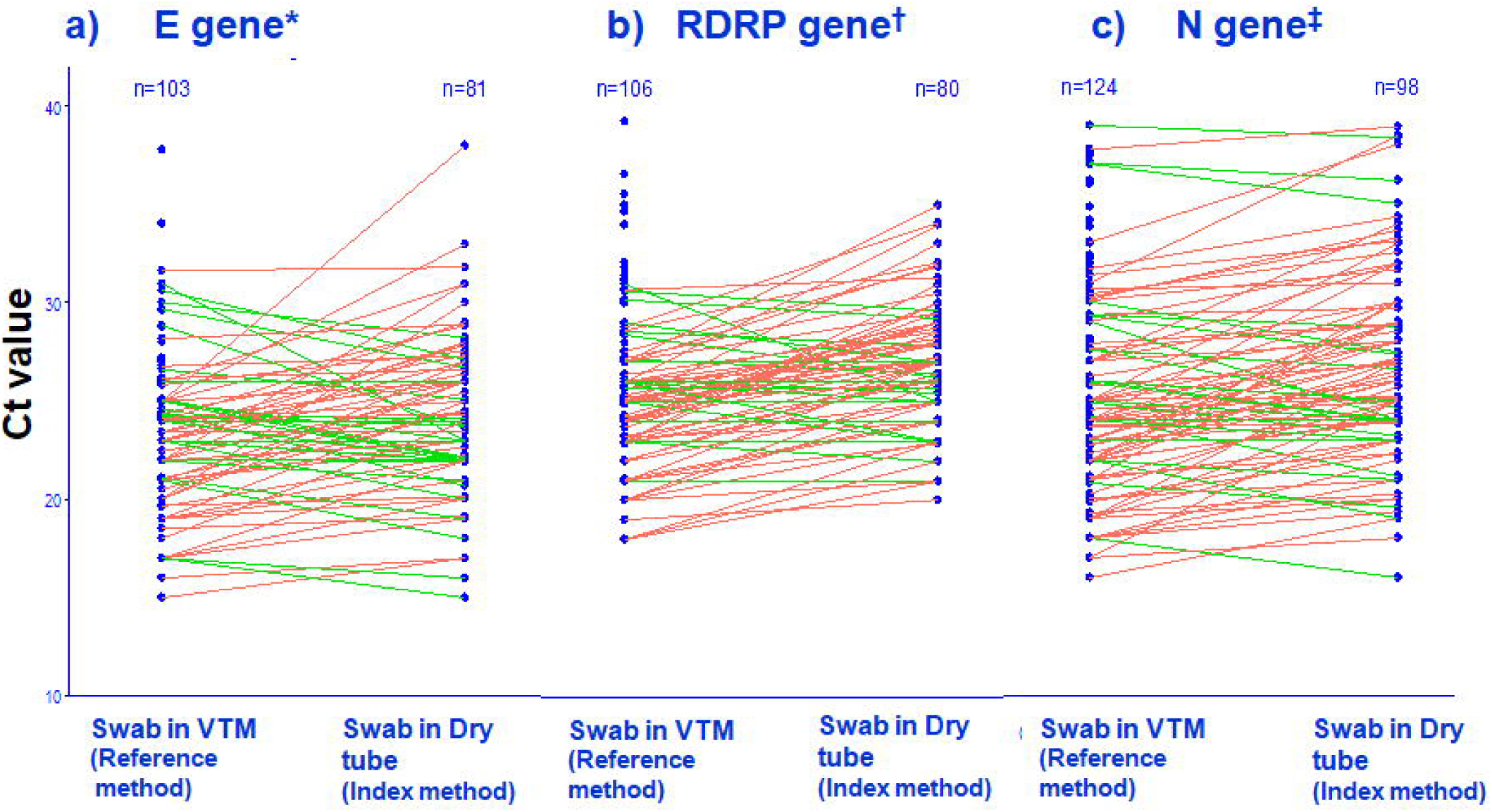
Flowchart showing enrolment of participants at two study sites (ICMR-NARI, Pune, Maharashtra and STM, Kolkata, West Bengal).

### Performance of index method

While 11% (128/1138) of the total clinic attendees tested positive for SARS-CoV-2 infection by the reference method, the index method involving transportation of swabs in dry tube environment detected 78% (101/128) of them thus reducing the overall detection to 8.8% (101/1138). Of the 584 samples tested at ICMR-NARI site, overall 9.6% samples (56/584) tested positive by reference method and 9.4% (55/584) were positive by index method. Of the 554 samples tested at STM, Kolkata, 72 (13%) were detected as positive by reference method and 55 (10%) by index method (Figure 4). The overall sensitivity of the index method was 78.9 % (95% CI 71% to 86%) and specificity was 99 % (95% CI 98% to 99.6%). The observed overall agreement between the index and reference method was 96.8 % and the discordance was 3.16 %; kappa value (k) was 0.83 (95% CI 0.77 to 0.89, SE=0.030). Site disaggregated data are presented in Table 3.

**Table 3.**
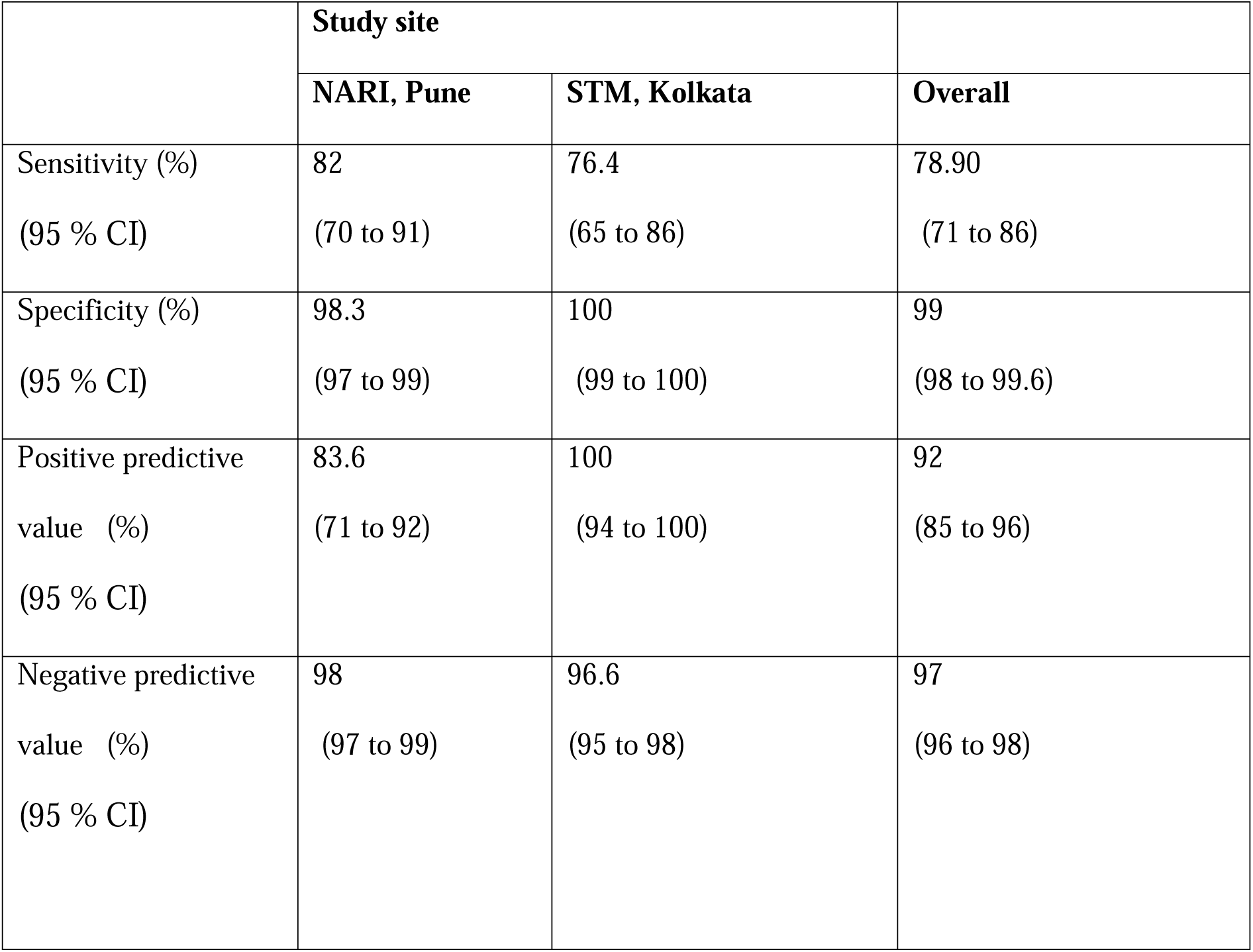
Performance of the RNA Extraction Free Index Method in Current Investigation

**Figure 4.**
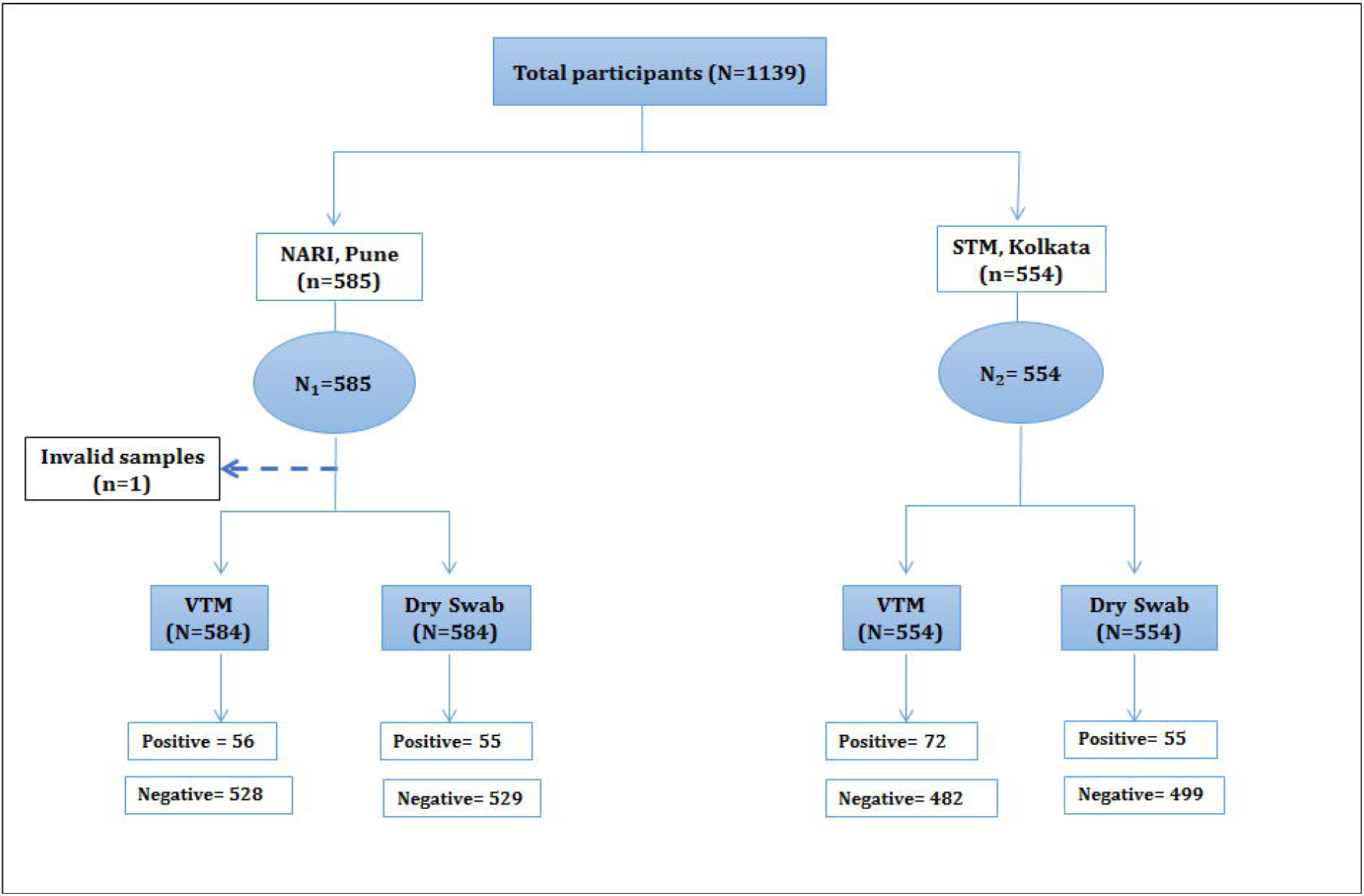
Scatter plot of Ct values for matched samples tested by reference and index method for a) E gene b) RdRP gene and c) N gene * Of 103 samples having E gene detection through VTM, 81 were detected through index method † Of 106 samples having RdRP gene detection through VTM, 80 were detected through index method ‡ Of 124 samples having N gene detection through VTM, 98 were detected through index method

## Discussion

Conducting research during outbreak situation faces many challenges. Lengthy start-up period before one could carry out observational research in pandemic situation has been cited as one of these challenges [22], and the other challenges are reactive approaches, socio-political pressures to approve repurposed or promising drugs [23] or diagnostic kits and urgency of the researchers to inform public health decisions. Besides prompt implementation against such background, the strength of the current investigation rests with its methodology. Firstly, a study population akin to the individuals, on which the index method could be applied in future, was assembled. Secondly, both the reference as well as index method pertaining to SARS-CoV-2 diagnosis were applied to all the study participants and laboratory investigators remained blinded to such assignments at both the study sites, which independently conducted their investigations.

With increasing demand for testing in pandemic situation, several researchers have explored the possibility of utilizing alternative specimen collection procedures, processing steps and testing methods. Direct heating of nasopharyngeal swab specimens in universal viral transport (UVT) medium at 65 °C for 10 minutes without RNA extraction reportedly yielded sensitivity comparable to the standard method [14]. On the contrary, an earlier evaluation of a direct extraction method using buffer eluates of the swabs transported in dry tube (without transport medium) and heat treatment at 98°C for 6 minutes against the reference method on 978 clinical samples yielded an overall sensitivity of 56 % (95% CI 49.8 % to 61.6 %) and specificity of 95 % (95% CI 93.4 % to 96.8 %) [20]. Pretreatment of such buffer eluates with Proteinase K followed by heat inactivation was found to improve sensitivity in a pilot laboratory assay of SARS-CoV-2 [24], consistent with the previous reports from other researchers [13].

We conducted the current assessment to evaluate a similar approach of direct extraction from dry-swabs using TE buffer and proteinase K followed by heat inactivation from naso-pharyngeal specimens collected from consecutive clinical attendees. This modification over an earlier version of the test approach [17] increased the overall sensitivity from 56% to 79%.

Contrastingly, Srivastan et al [25] reported much higher sensitivity (100%) and specificity (99.4%) with direct extraction from dry-swabs using low-TE buffer elution, proteinase K pre-treatment and heat inactivation. Noticeably, the study by Srivastan et al., used archived samples as well as anterior nasal dry-swabs collected as convenience specimens. Such designs are prone to introduction of biases that we could avoid by enrolling consecutive clinic attendees from two different clinic settings.

Heat map-based visualization, in the present investigation, demonstrated that the standard RNA extraction method exhibited enhanced detection of gene targets with lower Ct values (corollary of higher RNA concentration or viral load in a given condition) compared to the dry swab elution where RNA-fragmentation during heat inactivation remains a possibility, which could lead to reduced sensitivity. The difference could further be explained by purification that takes place during RNA extraction. Moreover, concentration of RNA that is achieved and removal of PCR inhibitory substances during RNA extraction also could contribute to better yield due to intact high-quality RNA available for RT-PCR. Noticeably, Chen et al (2020) reported that there was a 50%-66 % drop in RNA copy number after heating at 80°C for 20□minutes [26] while different inactivation methods were compared. In the present study, the index method failed to detect 17 % and 23% of positive specimens at ICMR-NARI site and STM, Kolkata site respectively.

Noticeably, eight positive specimens with low Ct values detected by the reference method were missed out by the extraction free method at STM, Kolkata. This could be due to the compromised quality of RNA during extraction process which failed to amplify all three viral genes. The low detection at STM was not related to site-specific performance issue as the internal control was detected at low Ct values in these specimens. Rather, different heating methods used at two study sites could explain the difference in recorded sensitivity due to resulting difference in time of exposure of samples to 98°C. Hasan et al, (2020) compared standard method with direct extraction method and showed enhanced detection of human RNase P compared to viral genes with a difference of 1-6 Ct values [14]. Burton and colleagues (2021) therefore recommend local validation of heat-inactivation and examination of its effects on molecular testing due to considerable variations observed in different studies [27]. These findings underline the importance of paying attention to the heat treatment method used while extraction free methods for viral diagnostics would be considered.

Importantly, Smyrlaki et al (2020) carried out extensive standardizations of different heat inactivation protocols. The authors reported that all high temperature (≥95 °C) conditions resulted in similar Ct values and recommended inactivation at 95°C to 98°C [16]. On the other hand, Mallmann et al (2021) tested different conditions and reported that pretreatment with Proteinase K and heat treatment at 98°C yielded best results with Ct values similar to that in standard method.

It was observed in the current investigation that the detection of SARS-CoV-2 N gene target was superior compared to E and RdRP genes. This is in agreement with the previous reports [28], where N-gene based RT-PCR was shown to be more sensitive due to relative abundance of N gene subgenomic mRNA [29]. The primer–probe set for N1 gene showed better performance due to shorter amplicon size in another heat inactivation protocol as well [16]. Hence, we maintain that the primer and probe sets should be carefully chosen if heat inactivation methods are to be deployed. Our study has further highlighted the importance of deploying appropriate heat treatment method if RNA-extraction-free detection technique is to be deployed.

In conclusion, the evaluated index-method has the potential to serve as an alternative protocol for identifying SARS-CoV-2 infection in resource limited settings. However, the following observations appear demanding if using this method in program setting is to be considered; a) requirement of carefully selected primer and probe sets for better outcome and b) the necessity of selecting appropriate heat treatment method. The lower sensitivity of this RNA-extraction-free RT-PCR method in real world setting appears to be one of its limitations.

## Data Availability

The data presented in this study are available on request from the corresponding author. The data are not publicly available due to ethical reasons.

## Acknowledgments

We thank Dr. Shailaja Bhavasar, Senior Medical officer, Bhosari Hospital, Dr. Vikalp Bhoi: Medical Officer, Bhosari Hospital, Ms Asawari Todewale: Laboratory Technician, Project, Mr. Mahibub Attar: Laboratory Assistant I, ICMR-NARI, Mr. Michael Pereira: Technical Officer-C, ICMR-NARI, Mr. Atul Sirsat: Technician-2, ICMR-NARI, Mrs. Dipali Kale: Senior Laboratory Technician-1, ICMR-NARI, Ms. Jyotsna Gokavi: Junior Research Fellowship, ICMR-NARI and COVID-19 diagnostic team at ICMR-NARI for helping in participant enrollment, sample collection and sample processing at ICMR-NARI site. We thank Dr Rinku Chakrabarti, Medical Officer and Dr Subhra Chattopadhyay, Senior Resident at STM, Kolkata for helping in laboratory assays at STM, Kolkata site.

## Ethical Statement

The authors assert that all procedures contributing to this work comply with the ethical standards of the national and institutional committees on human experimentation and with the Helsinki Declaration of 1975, as revised in 2008. The study was approved by the ICMR Central Ethics Committee on Human Research (CECHR) (CECHR 018/2020 dated 11^th^ August 2020 and 24^th^ October 2020). The study was also approved by the local Institutional Ethics Committees of two study sites –Ethics Committee, ICMR National AIDS Research Institute (NARI), Pune (Ref NARI/EC/Approval/20-21/385 dated 14^th^ August 2020) and Clinical Research Ethics Committee (CREC-STM), School of Tropical Medicine, Kolkata (Rf No. CREC-STM/613 dated 10^th^ August 2020).

## Author Contributions

SP conceived and designed the study, took part in analysis and was the technical advisor and national coordinator; MJ and SA were the study investigators; MJ and KJS contributed to data analysis, interpretation of data along with graphical representation; SG, AM, BC, BB, AR MT, AA, VS, BBa, BG, SD and AC were site investigators and were responsible for clinical, epidemiologic and laboratory investigations at the respective assessment study sites; RMP contributed to technical review of the study; BBh participated in discussion around development of the study protocol and provided critical inputs during implementation of the evaluation work and in drafting of the reports; MJ, SA,VS, BB and JS wrote the first draft of the manuscript. Further, SP revised and critically reviewed the manuscript. All authors read and approved the final version.

## References

1. Andrew Joseph. (2020). WHO declares coronavirus outbreak a global health emergency; 30 Jan 2020. https://www.statnews.com/2020/01/30/who-declares-coronavirus-outbreak-a-global-health-emergency/. Accessed on 10 July 2021.

2. WHO (2020), WHO Director-General’s opening remarks at the media briefing on COVID-19; 11 March 2020. (https://www.who.int/director-general/speeches/detail/who-director-general-s-opening-remarks-at-the-media-briefing-on-covid-1911-march-2020). Accessed on 10 July 2021.

3. Vaughan WT. (1921). Influenza: An epidemiologic study. The American Journal of Hygeine, Monograph series No.1. New Era printing company, Lancaster. Available from: (http://hdl.handle.net/2027/nnc2.ark:/13960/t6qz30017). Accessed on 10 July 2021.

4. Panda S, et al. (2021). Face mask - An essential armour in the fight of India against COVID-19. Indian J Med Res; 153: 233–237.

5. Mina MJ, Andersen KG. (2021) COVID-19 testing: One size does not fit all. Science; 371: 126–127.

6. Caruana G, et al. (2020) Diagnostic strategies for SARS-CoV-2 infection and interpretation of microbiological results. Clin Microbiol Infect; 26: 1178–1182.

7. Lu X, et al. (2020) US CDC Real-Time Reverse Transcription PCR Panel for Detection of Severe Acute Respiratory Syndrome Coronavirus 2. Emerg Infect Dis; 26: 1654–1665.

8. Tang YW, et al. (2020). Laboratory Diagnosis of COVID-19: Current Issues and Challenges. J Clin Microbiol; 58: e00512–20.

9. Corman VM, et al. (2020) Detection of 2019 novel coronavirus (2019-nCoV) by real-time RT-PCR. Euro Surveill; 25: 2000045.

10. Nalla AK, et al. (2020) Comparative Performance of SARS-CoV-2 Detection Assays Using Seven Different Primer-Probe Sets and One Assay Kit. J Clin Microbiol; 58: e00557–20.

11. Calvez R, et al. (2020) Molecular detection of SARS-CoV-2 using a reagent-free approach. PLoS One; 15: e0243266.

12. Alcoba-Florez J, et al. (2020) Fast SARS-CoV-2 detection by RT-qPCR in preheated nasopharyngeal swab samples. Int J Infect Dis; 97: 66–68.

13. Chu AW, et al. (2020) Evaluation of simple nucleic acid extraction methods for the detection of SARS-CoV-2 in nasopharyngeal and saliva specimens during global shortage of extraction kits. J Clin Virol ; 129: 104519.

14. Hasan MR, et al. (2020) Detection of SARS-CoV-2 RNA by direct RT-qPCR on nasopharyngeal specimens without extraction of viral RNA. PLoS One; 15: e0236564.

15. Fomsgaard AS, Rosenstierne MW. (2020) An alternative workflow for molecular detection of SARS-CoV-2 - escape from the NA extraction kit-shortage, Copenhagen, Denmark, March 2020. Euro Surveill ; 25: 2000398.

16. Smyrlaki I, et al. (2020) Massive and rapid COVID-19 testing is feasible by extraction-free SARS-CoV-2 RT-PCR. Nat Commun ; 11: 4812.

17. Kiran U, et al. (2020) Easing diagnosis and pushing the detection limits of SARS-CoV-2. Biol Methods Protoc ; 5: bpaa017.

18. de Paula VS, Villar LM, Coimbra Gaspar AM. (2003) Comparison of four extraction methods to detect hepatitis A virus RNA in serum and stool samples. Braz J Infect Dis ; 7: 135–141.

19. Specimen collection, transport and packaging guidelines for 2019 novel Coronavirus (2019-nCoV). https://www.mohfw.gov.in/pdf/5Sample%20collection_packaging%20%202019-nCoV.pdf

20. Indian Council of Medical Research (ICMR). Validation of a dry swab based sample collection and RNA-extraction-free diagnostic method for SARS-CoV-2: A multicentric study. ICMR Report, September 2020.

21. Flahault A, Cadilhac M, Thomas G. (2005) Sample size calculation should be performed for design accuracy in diagnostic test studies. J Clin Epidemiol ; 58: 859–862.

22. Rishu AH, et al. (2017) Time required to initiate outbreak and pandemic observational research. J Crit Care ; 40: 7–10.

23. Ueda M, et al. (2021) Japan’s Drug Regulation During the COVID-19 Pandemic: Lessons From a Case Study of Favipiravir. Clin Pharmacol Ther, doi: 10.1002/cpt.2251.

24. Mallmann L, et al. (2021) Proteinase K treatment in absence of RNA isolation classical procedures is a quick and cheaper alternative for SARS-CoV-2 molecular detection. J Virol Methods : 293:114131.

25. Srivatsan S, et al. (2020) SwabExpress: An end-to-end protocol for extraction-free COVID-19 testing. bioRxiv: the preprint server for biology, 2020.04.22.056283. https://doi.org/10.1101/2020.04.22.056283.

26. Chen H, et al. (2020) Influence of Different Inactivation Methods on Severe Acute Respiratory Syndrome Coronavirus 2 RNA Copy Number. J Clin Microbiol ; 58: e00958–20.

27. Burton J, et al. (2021) The effect of heat-treatment on SARS-CoV-2 viability and detection. J Virol Methods ; 290:114087.

28. Chu DKW, et al. (2020) Molecular diagnosis of a novel coronavirus (2019-nCoV) causing an outbreak of pneumonia. Clin Chem ; 66: 549–555.

29. Moreno JL, et al. (2008) Identification of a coronavirus transcription enhancer. J Virol ; 82: 3882–3893.

